# Responses to COVID-19 Threats: An Evolutionary Psychological Analysis

**DOI:** 10.1101/2022.06.20.22276460

**Authors:** Stephen M. Colarelli, Tyler J. Mirando, Kyunghee Han, Norman P. Li, Carter Vespi, Katherine A. Klein, Charles P. Fales

## Abstract

Responses to COVID-19 public health interventions have been lukewarm. For example, only 64% of the US population has received at least two vaccinations. Because most public health interventions require people to behave in ways that are evolutionarily novel and mismatched with evolved human perceptual and decision-making mechanisms, it is imperative that we gain a better understanding of how people respond to public health information, including how they respond under different pandemic conditions and how specific groups may differ in their responses. We conducted two studies, using data from primarily public sources. We found that state-level COVID-19 threats (e.g., infection and mortality rates) had no relationships with mental health symptoms, suggesting that people were not attending to threat information. This result is consistent with the evolutionary psychological explanation that COVID-19 threat information is insufficient to activate the human behavioral immune system. Furthermore, individual differences affected how people responded to COVID-19 threats, supporting a niche picking explanation. Finally, aggregate state IQ levels correlated positively with aggregate vaccination rates, suggesting that intelligence can partially counteract the evolutionary novelty of abstract threat information, supporting the savanna-IQ interaction hypothesis. We conclude with policy implications for improving interventions and promoting greater compliance.

---

The number of deaths in the U.S. from the COVID-19 pandemic (over one million) has exceeded the U.S. total from the Spanish Influenza epidemic (Curley, 2021), which had been the deadliest pandemic in U.S. history (Barry, 2020).^1^ Worldwide, over six million people have died from COVID-19. While there have been significant improvements in scientific and public understanding of the disease, progress with public health interventions remain disappointing (Cummins, 2022; Ishak, 2022; Lewis, 2021; Nan et al., 2022; Smith et al., 2022). For example, despite the severity of the COVID-19 pandemic and the widespread availability of safe and effective vaccines (the best-known way to defeat the pandemic), only 63% of the U.S. population was fully vaccinated at the beginning of 2022 (Center for Disease Control and Prevention, 2022). In Hong Kong, which experienced a COVID-19 surge during January - March 2022, only 64% of its population was vaccinated and only 47% among the elderly (Smith et al., 2022). Throughout the world, only 59% of the population has been vaccinated with at least two doses (Ritchie et al., 2021). These rates are much lower than for other serious infectious diseases. For example, approximately 83% of the world population has been vaccinated for polio, DPT, and measles (Muhoza et al., 2021).

Although the COVID-19 pandemic will likely run its course, there will be other pandemics (Olshaker & Osterholm, 2017). Moreover, given the unprecedented scale of global travel, future pandemics have the potential to be as widespread and severe as (if not more so) than the COVID-19 pandemic. Thus, it is imperative that we gain a better understanding of how people respond to public health information, how they respond under different pandemic conditions, and how specific groups may differ in their responses. This paper has three purposes. First, we discuss three perspectives on how people may respond to COVID-19 threats: (a) Responding to differential threat information, (b) ignoring threat information, and (c) responding based on individual differences. Second, we present results of two studies to provide evidence on these perspectives. Finally, we argue that an evolutionary psychological perspective can be helpful for understanding how people respond to COVID-19 (and other pandemic) threats and for developing effective behavioral public health interventions.

## Responding to Differential Threat Information

One view of how people respond to public health information and directives is that they, as rational thinkers (e.g., Cushman, 2020), gather available information, assess their situation – particularly their local situation – and respond appropriately. During the first phase of the pandemic in the U.S. (March – May 2020), infection and mortality rates varied by geography, with higher infection rates on the coasts and in larger population centers (CDC COVID-19 Response Team, 2020). If people were responding to differential threat conditions, reactions should differ by threat environments. For example, where local threat levels were low, we would expect less caution, less concern about becoming infected, and less anxiety. On the other hand, where local threat levels were high, there should be more caution, more concern about health risks, and greater levels of anxiety.

Unfortunately, understanding of the disease was spotty and variable during Phase I (∼January - June 2020) (Lewis, 2021). It was still unclear how infectious the coronavirus (SARS-CoV-2) was, with RO index (a measure of contagion) estimates ranging from 1.6 to 6.5 (Achaiah, Subbarajasetty, & Shetty, 2020). Some epidemiologists argued that high rates of infection were most likely to occur only in populated urban areas (CDC COVID-19 Response Team, 2020). It was also unclear whether infected individuals transmitted the disease prior to showing symptoms or only after symptoms appeared (Slifka & Gao, 2020; World Health Organization, 2020). Messages about wearing masks varied from unnecessary to a good idea to essential (Eikenberry et al., 2020; Worby & Chang, 2020). It was unclear who would be most susceptible to the virus in terms of infection, morbidity, and mortality. Effective vaccines were not available until over a year from the time the pandemic started, limiting public health interventions to keeping the public informed of infection and mortality rates and imposing lockdowns to prevent the spread of the virus. Therefore, standard measures for assessing responses to public health directives (e.g., vaccination and mask wearing rates) were not available or feasible.

Mental health symptoms were one of the few – albeit indirect – indicators of how people were responding to the pandemic and to public health information (Cullen et al., 2020; McGinty, 2020; Pfefferbaum & North, 2020). For example, increases in anxiety could reflect attention to infection and mortality statistics as well as concerns for safety and well-being. Increases in depression and loneliness could reflect the effects of isolation from lockdowns. It would seem reasonable to expect that people, especially in areas with high infection and mortality rates, would show higher rates of anxiety, while those with stricter lockdown requirements might show greater levels of depression and loneliness. Indeed, several studies found evidence for decreases in mental health during the first phase of the pandemic (González-Sanguino et al., 2020; McGinty et al., 2020). However, other studies found that the pandemic had minimal to no effects on psychological well-being. For example, a study conducted in Germany found that life satisfaction and positive affect barely decreased between March and May 2020 (Zacher & Rudolph, 2020). Luchetti and colleagues (2020) found, in a nationwide sample of U.S. adults, no mean changes in loneliness across January/early February 2020, late March, and late April 2020.

One explanation for the mixed results of these studies is that they sampled populations facing different threat levels. If threat levels varied across samples – a likely possibility – then people facing greater threats would exhibit greater mental health problems. This would suggest that people were responding to differential threat information. Where infection and mortality rates were modest, people should exhibit less distress; where pandemic threats were the highest, people should experience the most distress. On the other hand, if mental health symptoms do not vary across samples, this would suggest that people were ignoring threat information.

## Ignoring Threat Information

Ignoring – or not responding to – local threat information is also a likely scenario given that our evolved threat detection system is mismatched with the modern realities of the COVID-19 pandemic (e.g., Li et al., 2018; Seitz et al., 2020). People typically respond to threats that are *immediate and obvious—*threats that are salient enough to activate threat-based psychological mechanisms. Over millennia, people evolved to respond to personal narratives, identifiable individuals, and tangible threats (Moore, 1996). As a result, they are less likely to respond to evolutionarily novel information, such as pandemic statistics and commentaries about global trends. Of course, from a public health perspective, people *should* pay attention to COVID-19 threat statistics and be concerned about them. However, many are unlikely to do so.^2^

In addition, there is a mismatch between what our behavioral immune system evolved to respond to and early COVID-19 symptoms (Seitz et al., 2020). Our primary system for detecting and avoiding pathogens is the behavioral immune system (Schaller & Duncan, 2007). It operates by triggering avoidance responses to animate and inanimate objects that – recurrently over our evolutionary history – had a high probability of carrying infectious pathogens. This system triggers an avoidance response through the emotion of disgust (Cepon-Robins et al., 2021; Oaten et al., 2009). Both animate and inanimate objects with *obvious* signs of carrying infectious pathogens trigger the disgust response. Examples include spoiled food, feces, cadavers, sick animals, and people with noticeable signs of illness (e.g., blemishes, pustules, vomiting, a runny nose, skin pallor, deformities). The disgust response is normally followed by avoidance. However, most people infected with the coronavirus are initially asymptomatic, and early symptoms are often not severe (resembling the common cold). By the time people are severely ill, many are out of view – isolated – at home or in a hospital. Thus, there are little or no available inputs to activate the evolved behavioral immune system. This has been the case in all phases of the pandemic thus far.^3^

One might counter that the media and social media are full of images related to the ravages of COVID-19—and that these images should activate the behavioral immune system (Schaller et al., 2010). While there are abundant images of overworked health care workers, intensive care units overflowing with COVID-19 patients, spikey balls representing the virus, and even coffins, there have been – for a variety of reasons, including medical privacy laws – few images of identifiable victims in the throes of contagious infection (Lewis, 2020). Moreover, the images that we do see (of intensive care units filled with COVID-19 patients, ventilators, and so forth) are *evolutionary novel*. These images are unlikely to activate the behavioral immune system in the way that evolved signs of ill health (e.g., vomit, runny nose, pallor, wheezing, and deformities) do. Common media images during polio epidemics showing crippled children would be more likely to trigger the behavioral immune system than those of an unseen COVID-19 patient in a hospital room. Thus, it is not surprising that people with *direct experience* with COVID-19 – either experiencing symptoms themselves or having a friend of relative who was sick with COVID-19 – are most likely to suffer negative mental health symptoms (González-Sanguino et al., 2020).

## Responding Selectively based on Individual Differences

An evolutionary psychological perspective also suggests that people would respond *selectively* to COVID-19 threats, based on individual differences.

### Personality

The theories of niche picking, reactive heritability, and frequency dependent selection explain how individual differences in personality can evolve and be adaptive. For example, neuroticism and anxiety continue to be widespread traits because people with these traits, over evolutionary history, probably were more likely to survive and reproduce by playing it safe. Other traits (e.g., extraversion, openness to experience), however, may continue to be widespread traits because of the adaptive value of pursuing risky strategies. Pursuing a “safe” strategy (e.g., hypervigilance to threat information) may be an adaptive response for people with anxious or neurotic personalities. These individuals would be most likely to suffer depression, loneliness, or anxiety in the face of imminent danger, which in turn would trigger caution and isolating behavior. In contrast, a risky strategy – ignoring threat information and carrying on normally – may be more adaptive for those who are unflappable or extraverted (Nettle, 2006).

### Age

People may also respond selectively depending on their age. Both the “respond to differential threat information” and “the individual difference” explanations predict that older people would be most responsive to COVID-19 threats. Towards the end of Phase I, the scientific evidence became stronger and clearer that the elderly were among the most susceptible to COVID-19 infections *and* most likely to die from the disease (Liu et al., 2020). Moreover, within a year vaccines became available and the elderly were among those given priority to them. This information would suggest, from a rational thinking perspective (e.g., Cushman, 2020), that the elderly would view their threat level as high and would act accordingly. Furthermore, as people age, they become more cautious and sensitive to threats because they become more frail and susceptible to injury and infection. Evolution designed a variety of psychological adaptations to be switched on and off throughout the lifespan (Buss, 2009) and these mechanisms would also motivate the elderly to be more receptive, on average, to health precautions. All these factors suggest that older people would be more responsive to COVID-19 threats than younger people, including having higher vaccination rates.^4^

### Intelligence

An additional evolutionary psychological hypothesis suggests that COVID-19 threats may be influenced by general intelligence. The savanna-IQ interaction hypothesis proposes that general intelligence evolved to allow individuals to overcome evolutionarily novel problems (Kanazawa, 2010). People of higher intelligence are better able to process evolutionarily novel information, such as statistics and news reports about an invisible disease. Thus, people of higher intelligence may be more thoughtful after being inundated by reports about COVID-19 and may be more apt to take precautionary responses, such as getting vaccinated.

### Ideology

Finally, how people perceive COVID-19 threats and respond to them has been influenced by ideology in the U.S. Ideology has become something of an honest signal of in-group-out-group membership (Conway et al., 2021). Because early symptoms are mild and severely ill patients are isolated, the likelihood increases that responses to COVID-19 will be influenced by other factors, such as beliefs and ideology. Conversely, when a threat is obvious and immediate (e.g., polio in the case of a more obvious disease threat or a tidal wave in the case of an immediate and obvious weather-based threat), ideologies are unlikely to have much effect on threat responses.

## The Current Studies

In Study 1, conducted during the early months of the COVID-19 pandemic in the U. S. (May, 2020), we examined the relationship between variation in state-level COVID-19 threats and mental health symptoms. We also examined the relationship between personality characteristics and mental health symptoms. In Study 2 we use available (state-level) data to examine the effects of age, intelligence, and ideology on responses to COVID-19 threats two years later (2022). We use the percentage of state-level votes for Donald Trump in the 2020 U.S. presidential election as a proxy variable for ideology.

## STUDY 1

### Method

In Study 1, 418 individuals (67% response-rate) across the U.S. were recruited during the third week of May 2020 using Amazon’s Mechanical Turk. However, only 291 participants representing 13 states (see Table 1) were included after removing states with less than 10 participants. The mean age of the sample was 37.76 years (*SD* = 10.98 years). Of the 291 participants, 54.0% were male, 45.7% were female, and 0.30% identified as other. The self-reported major racial/ethnic composition of the sample was 68.4% Caucasian, 18.2% Black, 10.0% Asian, 6.5% Hispanic/Latino, 3.4% Native American, 0.7% Middle Eastern, 0.7% Hawaiian, and 0.3% Other. Most of the participants were married (49.1%), while 27.5% were single, 16.5% were in a relationship/not married, 5.5% were divorced/separated, and 1.4% were widowed. Education categories for the sample were as follows: high school degree or equivalent (6.5%), some college but no degree (11.7%), associate degree (11.0%), bachelor’s degree (52.9%), and graduate degree (17.9%).

**Table 1.** Sample Size, Mean (Standard Deviation) of Loneliness, Anxiety, and Depression by State

| State | *n* | Loneliness | Anxiety | Depression |
| --- | --- | --- | --- | --- |
|  |  | *M (SD)* | *M (SD)* | *M (SD)* |
| California | 55 | 2.39 (0.88) | 1.86 (0.72) | 1.72 (0.79) |
| Florida | 29 | 2.28 (0.87) | 1.76 (0.64) | 1.53 (0.65) |
| Georgia | 19 | 2.39 (0.53) | 1.75 (0.67) | 1.43 (0.68) |
| Illinois | 18 | 2.47 (0.91) | 1.93 (0.80) | 1.77 (0.73) |
| Michigan | 10 | 2.20 (0.67) | 1.77 (0.95) | 1.39 (0.49) |
| New York | 32 | 2.54 (0.85) | 1.93 (0.71) | 1.65 (0.81) |
| North Carolina | 20 | 2.21 (1.06) | 1.88 (0.93) | 1.63 (0.80) |
| Ohio | 17 | 2.65 (0.94) | 1.81 (0.51) | 1.67 (0.72) |
| Pennsylvania | 16 | 2.65 (0.98) | 2.09 (0.66) | 1.78 (0.72) |
| Texas | 40 | 2.40 (0.76) | 1.72 (0.68) | 1.63 (0.74) |
| Virginia | 15 | 1.99 (0.70) | 1.37 (0.46) | 1.44 (0.67) |
| Washington | 10 | 2.66 (0.45) | 2.03 (0.64) | 1.84 (0.78) |
| Wisconsin | 10 | 2.76 (0.55) | 1.98 (0.59) | 1.86 (0.82) |

*Note.* Loneliness (1-5 scale); Anxiety (1-4 scale); Depression (1-4 scale).

We assessed state-level COVID-19 indicators with archival data from the COVID-19 Tracking Project at *The Atlantic* (The COVID-19 Tracking Project at *The Atlantic*, 2020). These data were gathered from state health websites across the nation. State-level infection and mortality rates were selected and divided by the population of each state. We also captured variations in length of government-imposed lockdowns between states using data from *USA Today* (2020). Data provided by *USA Today* were based on official reports of state-level lockdowns and aggregated by Safe Graph (a California-based firm).

For mental health indicators, we measured loneliness with *The Loneliness Scale* (De Jong Gierveld & Van Tilburg, 1999), an 11-item Likert-style questionnaire (α = .83), depression severity with the 9-item *Patient Health Questionnaire* (α = .94; Kroenke & Spitzer, 2002), and state-anxiety with the 5-item shortened version of *The State-Trait Inventory for Cognitive and Somatic Anxiety* (α = .88; Marteau & Bekker, 1992). We measured the personality traits of Neuroticism (α = .73), Extraversion (α = .79), Conscientiousness (α = .70), Openness (α = .79), and Agreeableness (α = .76), with the MINI-IPIP (Donnellan et al., 2006).

### Results and Discussion

Table 1 presents the means and standard deviations of mental health variables (loneliness, anxiety, and depression) for 13 states. The lowest and highest means across states for each mental health variable were as follows: 1.99 (Virginia) to Wisconsin (2.76) for loneliness; 1.37 (Virginia) to Pennsylvania (2.09) for anxiety; 1.39 (Michigan) to Wisconsin (1.86) for depression. Table 2 presents correlations among state-level COVID-19 threats (number of days locked down, infection rate, and mortality rate) and mental health symptoms (loneliness, anxiety, and depression). The number of days locked down was not significantly related to any of the outcome variables (*r* = -.01 - .03). The mental health symptoms were not significantly related to either infection (*r* = .00 - .06) or mortality rates (*r* = -.00 - .07).

**Table 2.** Table 1. Correlations Among State-Level COVID-19 Threats, Mental Health Outcomes, and Personality

|  |  | Loneliness | Anxiety | Depression |
| --- | --- | --- | --- | --- |
| State-level indicators of COVID-19<sup>a</sup> | Number of days locked down | -.01 | -.00 | .03 |
|  | Infection rate | .06 | .06 | .00 |
|  | Mortality rate | .06 | .07 | -.00 |
| Personality Traits<sup>b</sup> | Neuroticism | .57 | .59 | .57 |
|  | Extraversion | -.18 | -.15 | -.12 |
|  | Openness | -.29 | -.23 | -.24 |
|  | Agreeableness | -.36 | -.23 | -.26 |
|  | Conscientiousness | -.41 | -.39 | -.42 |

<sup>a</sup> $n = 291$ $p > .05$ for all $rs$ .
<sup>b</sup> $n = 418$ ; $p < .05$ for all $rs$ .

Figure 1 shows mean scores of loneliness, anxiety, and depression across 13 states. States are listed in an ascending order of days of lockdown. Georgia had the shortest lockdown length (27 days), whereas Michigan had the longest lockdown (73 days). We expected an upward monotonic trend if days of lockdown were positively related to loneliness. However, no linear pattern was observed, suggesting that loneliness mean scores were not associated with the length of lockdowns. Similar results were found for infection and mortality rates. Although mean mental health outcome scores did not vary in a meaningful way across states, personality traits were associated with loneliness, depression, and anxiety (see Table 2) (*r* = -.42 to .59, *p* < .05).

**Figure 1.**
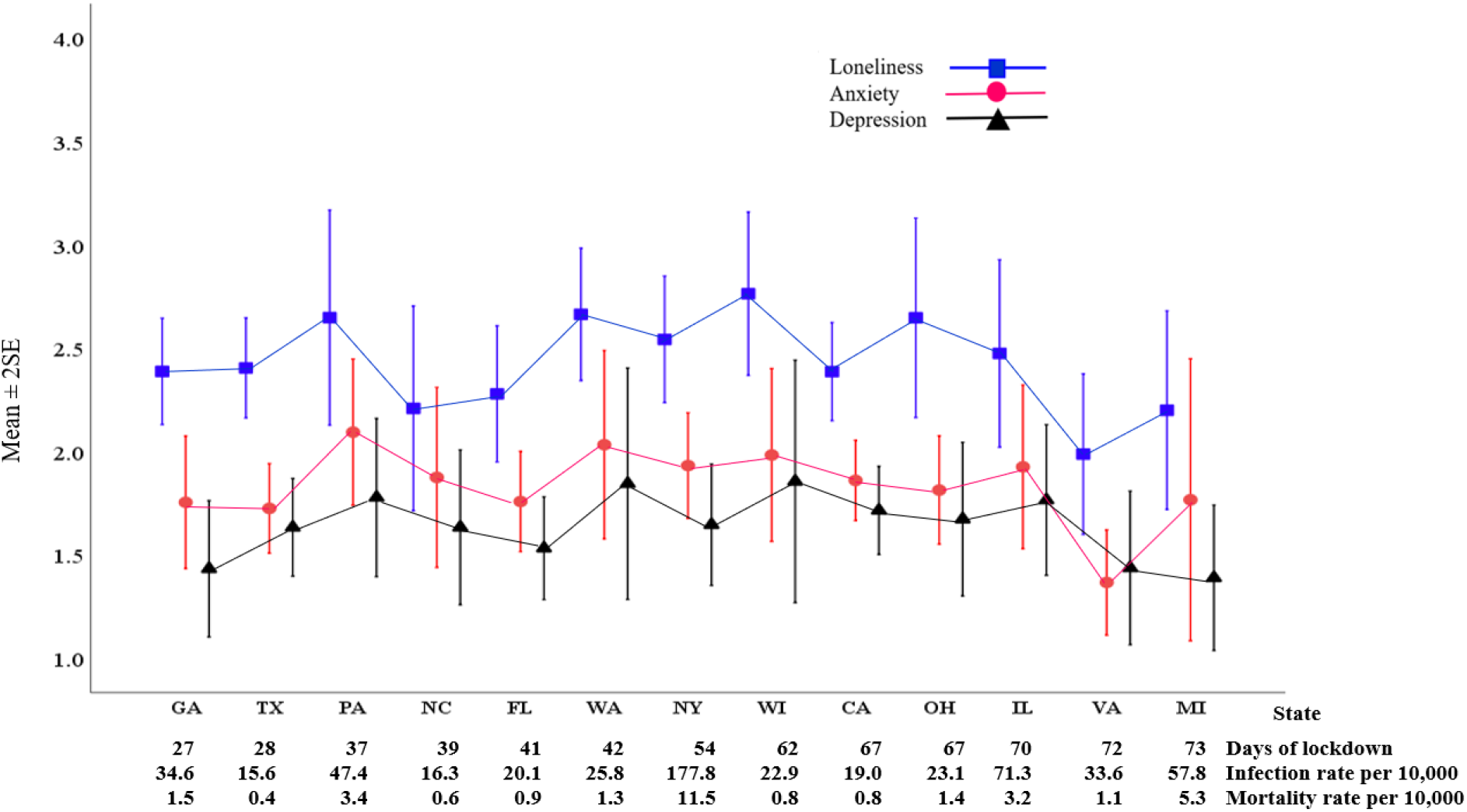
Mean Mental Health Outcome Scores by Days of Lockdown, Infection Rate, and Mortality Rate

Our findings suggesting no association between threats of COVID-19 and mental health symptoms are similar to findings in other studies examining objective measures of COVID-19 threats and mental health (Nocentini et al., 2021). As we argued, people may not be attending to or believe implications of infection and mortality rates from COVID-19. However, our results also suggest that some people may be more sensitive to real or imagined health risks (Jungmann & Witthöft, 2020). We found that people low in emotional stability (i.e., high in neuroticism) reported greater negative mental health symptoms, while those with higher levels of extraversion, agreeableness, conscientiousness, and openness to experience reported greater levels of psychological well-being.

## STUDY 2

### Method

Current total vaccination rates for each U.S. state (as of February 6^th^, 2022) were gathered from the Mayo Clinic’s website (Mayo Clinic, 2022). The Mayo Clinic is a nonprofit hospital system and academic medical center that provides esteemed, often publicly-accessible medical research. We collected vaccination rates by age group (ages 5-11, 12-17, 18-64, and 65+) for each U.S. state from the Mayo Clinic website and the average vaccination rate for each age group was calculated.

We collected IQ data for each state from the World Population Review website (World Population Review, 2022). The World Population Review is an independent organization that seeks to provide normally inaccessible demographic data for public examination and use. For the data used in the present study, the World Population Review used a study conducted by the *Washington Post* that aggregated various measures of cognitive ability (IQ scores, SAT and ACT scores, and education level) into an overall IQ measure for each U.S. state. Data pertaining to 2020 presidential election results were drawn from CNN’s website (CNN, 2020), which tracked which states were won by Joe Biden or Donald Trump, as well as the percentage of the votes going to either of the two candidates for each state.

### Results

The mean full vaccination rate for each age group across all U.S. states is shown in Table 3. Vaccination rates increased with age, with the youngest age group (5-11) showing the lowest percent vaccinated (*M* = 21.89, *SD* = 9.60) and the oldest age group (65+) showing the largest percent vaccinated (*M* = 94.08, *SD* = 4.65). The 65+ age group also showed the least amount of variability in vaccination (*SD* = 4.65) of all the age groups.

**Table 3.**
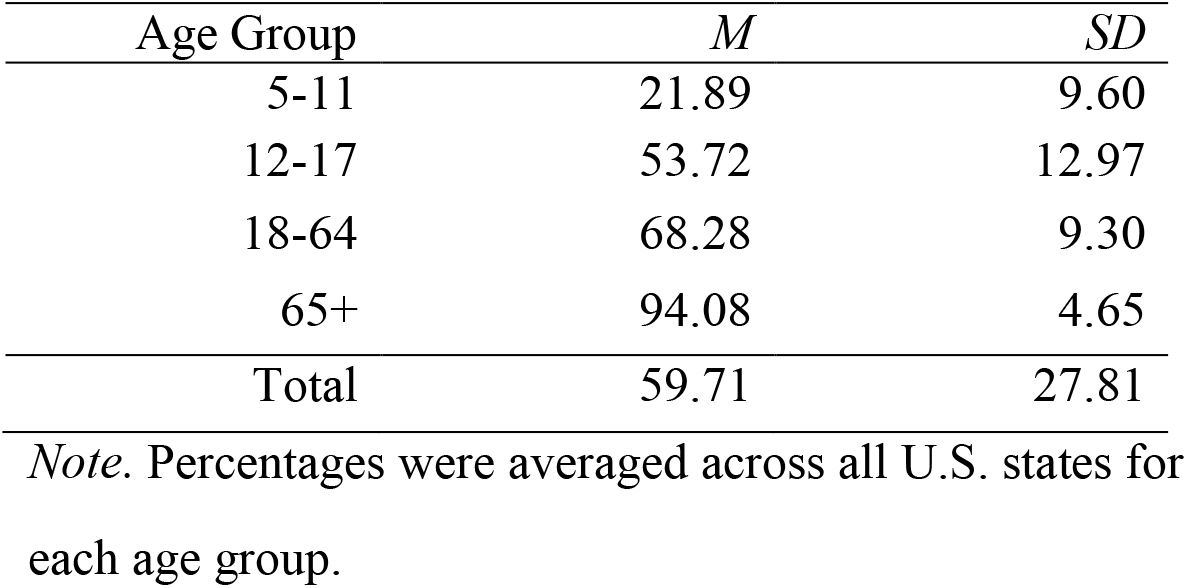
Percent of U.S. Population Fully Vaccinated by Age Group

The relationship between the total vaccination rates of U.S. states and each state’s average IQ was tested, along with the relationship between states’ total vaccination rates and the total percentage of each state’s vote that went to Trump in the 2020 presidential election. There was a significant positive correlation between the percentage of the population fully vaccinated and average IQ across all states (*r* = .35, *p* < .001). There was also a significant and strong negative correlation between total vaccination rate and the percentage of the vote that went to Trump across all states (*r* = -.88, *p* < .001).^5^

Like the results in Study 1, these results also show that individual differences influence how people responded to COVID-19 pandemic information. Both age and intelligence correlated positively with vaccination rates, while support for Donald Trump in the 2020 presidential election correlated negatively with vaccination rates.

### General Discussion

We found that state-level COVID-19 threats (number of days locked down, infection rate, and mortality rate) had no relationships with mental health symptoms (loneliness, anxiety, and depression) during the early months of the pandemic. Mean scores of mental health variables did not vary in a meaningful way across states. That large numbers of people were not experiencing negative mental health symptoms may be why so many continued to ignore preventative guidelines—despite a continuous stream of information about objective threats, including multiple waves of infections and deaths. Our finding of a lack of association between COVID-19 threat exposure and mental health is consistent with other studies examining objective measures of COVID-19 threats and mental health (Nocentini et al., 2021). This suggests that many people are neither attending to nor responding to COVID-19 threat levels. These findings are consistent with the two evolutionary psychological explanations. First, the early symptomology (i.e., lack of visual cues) of COVID-19 and how patients are cared for (in isolation) are unlikely to activate our behavioral immune system. Second, people tend not to respond instinctively and actively to abstract information (such as infection and mortality statistics)—as they would with visual cues of or narratives about individual people (Li et al., 2018).

However, individual differences were associated with how people responded to COVID-19 threats. Of the Big Five personality characteristics, neuroticism correlated strongly with negative mental health symptoms (loneliness, anxiety, and depression), while all of the other traits correlated negatively with adverse mental health symptoms. While our data are cross sectional, these correlations suggest some impact of the pandemic in that they are stronger than typical correlations among the Big Five and negative mental health outcomes prior to the pandemic (e.g., Bunevicius, Katkute, & Bunevicius, 2008). The results from Study 2 also found that individual differences played an important role in how people responded to COVID-19 threats. Two years after the outbreak of the pandemic and after vaccines were available, the elderly (65+) had considerably higher rates of being fully vaccinated (94%) than all other age groups.

Finally, aggregate state IQ levels correlated positively with aggregate vaccination rates. Thus, although people do not possess psychological mechanisms enabling them to respond instinctively and actively to abstract information (such as infection and mortality statistics), intelligence (which is correlated with education and scientific literacy) can, to some extent, counteract this. These results are compatible with other studies that found people who accept the tenets of modern science were most likely to follow public health recommendations during the pandemic (Brzezinski et al., 2020) and that educational level is negatively associated with vaccine hesitancy (Khubchandani et al., 2021; Killgore et al., 2021).

## Limitations

A limitation of Study 1 is that the mismatch implication – people not having evolved to respond to novel stimuli such as threat information – was supported by the absence rather than the presence of significant results. Moreover, the lack of correlations between COVID-19 threat indicators and mental health could be due to many unidentified factors. Thus, while we contribute to the literature by outlining a potentially strong explanation for a pressing, real-world phenomenon, we only provide indirect empirical support for the mismatch hypothesis. More rigorous tests—including experimental methods that manipulate different ways of conveying the virus (e.g., evolutionarily novel statistical reporting versus visual presentation of severe outcomes) are clearly needed to substantiate the hypotheses. Another promising route is to investigate moderators that may influence when the correlation between COVID-19 and mental health, as well as vaccination and other precautions, becomes significant.

Another possible limitation is the use of state-level data. While it would have been preferable that all data were at the individual level, certain types of data relevant to our research questions (e.g., matching individual vaccination status with IQ, who an individual voted for) would be restricted and possibly inaccurate. Nevertheless, we believe that our results are broadly indicative of the relationships we assessed. We recommend that more granular research should be conducted in future studies to delve further into these relationships.

## Implications

Despite the mitigating influence of scientific literacy and education, the remediation of modern, global pandemics through public health interventions is and will remain difficult. Most public health interventions require people to behave in ways that are evolutionarily novel and mismatched with evolved human perceptual and decision-making mechanisms. This includes understanding and accepting abstract scientific information, avoiding or staying distant from people who do not seem ill, staying at home when feeling fine, wearing face coverings, and getting injected with foreign substances. The greater the degree that a desired behavior is at odds with its adaptive value over millennia of human evolution, the more difficult it will be for an intervention to effectively encourage that behavior (Jones, 2001). For example, because frequent social interaction with friends and family has been adaptive to humans for millennia, people will be more resistant to public health interventions that restrict normal human interaction (lockdowns, social distancing, wearing face masks) than to interventions that facilitate social interaction.

Our findings, combined with the above evolutionary logic, have four major implications for public policy. First, expecting broad voluntary compliance – especially during COVID-19-like pandemics – is unrealistic. For the majority of people, some mandatory regulations may be necessary to assure sufficient compliance. This can occur through mandates from government or other institutions, such as employers. Typically, countries with stronger vaccine mandates and social pressure for vaccination have higher vaccination rates (Suliman et al., 2021). Interventions that link compliance with valued evolutionary-based rewards (such as status, access to status, or mating opportunities) are more likely to be successful. For example, making admission to workplaces, schools, and gathering places for singles contingent upon wearing masks or having proof of vaccination is likely to increase compliance.

Second, because people respond selectively to pandemic threats based on individual differences, communication strategies should be selectively tailored to specific groups. People who are most likely to be affected by a pandemic – the elderly in the case of COVID-19 and parents of young children in the case of polio – are more likely to use effortful appraisal—what Kahneman (2011) calls System 2 thinking and what others have referred to as systematic or central processing (Petty & Cacioppo, 1980). Thus, information and appeals to the most vulnerable groups should be designed to engage more elaborate processing, such as the presentation of high-quality and accessible scientifically backed arguments. Groups that consider themselves to be less vulnerable – and thus are less motivated to carefully process information – may be persuaded by more superficial methods such as using attractive celebrity endorsements (Petty & Cacioppo, 1984).

Third, responses to public health information can be tailored to the degree to which the disease is likely to activate the behavioral immune system. In the case of COVID-19, more work should be done on examining appropriate and believable imagery in public health communications. With other diseases with obvious harmful effects, as was the case with polio and its obvious crippling and deforming effects on children, the behavioral immune system would clearly be an ally in public health communications.

Finally, it must be acknowledged that belief systems are hard to change. If people believe that a vaccination is unsafe or that the negative effects of the disease are overstated—it will be difficult to change those beliefs with a rational argument based on scientific evidence. People use reason to find justifications for their beliefs, which in turn enhance their reputations within specific groups—not to find the true state of affairs (Mercier & Sperber, 2017; Yong, Li, & Kanazawa, 2021). Thus, to overcome opposition to public health policies, clearer explanations of their scientific basis are unlikely to be effective. A better strategy to get through to a skeptical public would be to use positive public health testimonials from high status individuals who are from those groups in which a majority of members are resistant to public health interventions.

In summary, the COVID-19 pandemic has generated a less than desirable response in places where people are relatively free to choose that response. We have provided an explanation based on evolutionary psychological principles and obtained empirical results consistent with this explanation and inconsistent with a more commonly accepted explanation. More work is needed, but findings from a growing number of studies indicate that a consideration of how the modern world is mismatched to how we have evolved to think, feel, and behave, can provide insights into the numerous problems that humans are now facing and why they are difficult to overcome.

## Data Availability

All data produced in the present study are available upon reasonable request to the authors.

## Footnotes

1 Severe acute respiratory syndrome coronavirus 2 (SARS-CoV-2) is the virus that causes COVID-19.

2 People responded pretty much the same during the Spanish Flu pandemic on the early 20^th^ century as they did now: ignoring public health recommendations to social distance and wear masks (Barry, 2020).

3 Contrast this with responses to the Polio epidemic. It was one of the most (if not *the* most) feared disease in the first half of the 20^th^ century. Unlike Covid-19, the symptoms of polio were obvious—paralysis and often death, primarily among infants and children (Baicus, 2012; Centers for Disease Control and Prevention [CDC], 2021). Prior to the vaccine and when the polio pandemic was at its worst (1948-1955), parents were extraordinarily cautious about letting their children go outside or to public gathering places (e.g., swimming pools), particularly in the summers, when the incidences of infection were highest (Mayo Clinic Staff, 2022).

With children, of course, vaccinations are parents’ decisions. We expect, though, that parents would follow a decision calculus based on the perceived threat of Covid to their children’s well-being. As the evidence became clear that young children were least susceptible to infection and least likely to become ill or die from Covid, we would expect that children would be the group least likely to be vaccinated. During the polio epidemic children were the most susceptible demographic group, and they were most likely to be vaccinated (Mayo Clinic Staff, 2022).

For elderly group, vaccination % ranged from 83.20 (Arkansas) to 99.90 (VT, RI, Main, WA, NH, MN, DE, WI) in 50 states. The mean % vaccination for states that voted for Biden was 96.67 (*SD* = 3.69), and for Trump it was 91.48 (*SD* = 4.05).

